# Neonatal platelet count trends during inhaled nitric oxide therapy

**DOI:** 10.1101/19007229

**Authors:** Christopher S Thom, Matthew Devine, Stacey Kleinman, Erik A Jensen, Michele P Lambert, Michael A Padula

## Abstract

Recent debate has focused on the significance of platelets generated in lung tissue. Here, we retrospectively analyzed platelet count changes in neonates requiring inhaled nitric oxide (iNO) pulmonary vasodilation therapy for pulmonary hypertension. There were no clinically significant changes in platelet count upon iNO initiation or during iNO therapy. Unexpectedly, platelet counts increased after iNO cessation. These findings argue against a clinically meaningful untapped pulmonary repository of megakaryocytes and platelets in this context, although acute platelet count increases might be expected after discontinuing iNO in some patients. Further work is needed to clarify the underlying etiology for these observations, and to better delineate the mechanisms for platelet count increases after recovery from lung injury.

## Introduction

Thrombocytopenia, platelet count below 150 ×10^9^ per L, results from inadequate production or increased consumption of platelets. Thrombocytopenia affects up to 35% of all NICU patients, including up to 90% of very low birth weight neonates (Gunnink *et al*, 2014). Donor-derived platelet transfusions are frequently given empirically to prevent bleeding in thrombocytopenic infants. The optimal platelet transfusion threshold is a point of contention, with wide clinical practice variations (Sparger *et al*, 2015). Importantly, platelet transfusions increase morbidity and mortality in preterm infants (Curley *et al*, 2018). These findings highlight a need to anticipate clinical scenarios that might permit more judicious use of platelet transfusions.

Traditionally, platelets have been thought to derive from precursor megakaryocytes in bone marrow (Elagib *et al*, 2018). However, some platelets are generated from megakaryocytes that have migrated to the lung (Lefrançais *et al*, 2017; Zucker-Franklin & Philipp, 2011). The clinical significance of lung-derived platelets remains a matter of debate.

Pulmonary arterial hypertension (PH) can severely diminish pulmonary blood flow and is frequently encountered in the neonatal intensive care unit (NICU). PH often results from persistent pulmonary hypertension of the newborn (PPHN), meconium aspiration syndrome, or abnormal lung development from congenital diaphragmatic hernia (CDH), omphalocele, or pulmonary hypoplasia. Acute severe neonatal PH can be treated with mechanical ventilation, direct pulmonary vasodilators such as inhaled nitric oxide (iNO), and/or extracorporeal membrane oxygenation (ECMO). PPHN and idiopathic PH are associated with thrombocytopenia (Mojadidi *et al*, 2014; Le *et al*, 2019).

We aimed to determine whether increased pulmonary blood flow in the setting of PH treatment affects platelet counts. Infants with severe PPHN have had diminished pulmonary circulation in their lifetime, with relatively naïve pulmonary vascular environments in which to study the effects of acute changes. Clinical scenarios in which neonates develop severe PH often include independently risk factors for thrombocytopenia, including sepsis (Guida *et al*, 2003). While this complicates parsing a clear association of PH and platelet counts, we reasoned that the rapid (seconds to minutes) effects of iNO would permit temporal correlation (Dupuy *et al*, 1992). In contrast, slow temporal changes in platelet counts are more likely derived from bone marrow production, as normally occurs over time during infancy (Sparger *et al*, 2015).

## Methods

### Subject identification

Subjects were identified via retrospective review of laboratory and clinical data from admissions to our neonatal intensive care unit (NICU) from 2010-2018. Inclusion criteria were admission at <28 d of age and receiving iNO. All infants that met these clinical inclusion criteria in our NICU would be expected to have clinically significant pulmonary hypertension. Indeed, most of these infants carried a formal diagnosis of pulmonary hypertension in their medical record. Excluded were subjects who received platelet transfusions or were treated with ECMO, as these are independent factors that would independently elevate or decrease platelet count, respectively. Those with diagnoses of CDH, pulmonary hypoplasia or omphalocele were excluded from our primary patient cohort. Those with CDH, who typically have abnormal lung parenchyma, were considered as a separate subpopulation.

To achieve adequate subject numbers without sacrificing specificity for the acute changes induced by iNO, we considered subjects for whom complete blood counts (CBCs) were available within 48h prior to iNO initiation and within 48h following iNO discontinuation. We also included results from time periods during iNO therapy, specifically within 48h after starting iNO and within 48h before stopping iNO. We focused on platelet counts, using red blood cell (RBC) and white blood cell (WBC) counts as control indices. We grouped up to 5 laboratory values within each time frame to obtain a best estimate of the average blood count values.

Respiratory severity score (RSS) correlates well with oxygenation index in neonates (Iyer & Mhanna, 2013). RSS is calculated by multiplying the fraction of inspired oxygen by mean airway pressure in mechanically ventilated subjects, and was used as a surrogate marker of pulmonary response to iNO. This was used in favor of oxygenation index, since in many instances the arterial partial pressure of oxygen was unknown.

### Statistical Analysis

GraphPad Prism 8 was used to calculate statistics and to generate graphs. Statistical significance was defined as a p-value less than 0.05 in all tests.

We analyzed laboratory values using mixed effects analysis and Tukey’s multiple comparison tests, given that some subjects did not have blood counts obtained in the analyzed time points during iNO therapy. Linear regression analysis determined best fit lines for temporal trends.

### Ethics

The Children’s Hospital of Philadelphia Institutional Review Board deemed this study exempt from review.

## Results

We retrospectively identified infants admitted to our quaternary care NICU at <28 d of age who were treated with iNO and had at least one complete blood count (CBC) performed in the 48h prior to initiating iNO *and* within 48h after stopping iNO (Table 1). The average duration of iNO therapy in this cohort was 22.7 d. We excluded those who required therapies that would confound endogenous platelet measurements, such as ECMO or allogeneic platelet transfusions. Our initial analysis also excluded patients with pulmonary hypoplasia, including those with omphalocele and congenital diaphragmatic hernia (CDH). We considered as a separate population those patients with CDH, to assess whether abnormal lung development might alter platelet count responses to iNO.

**Table 1.**
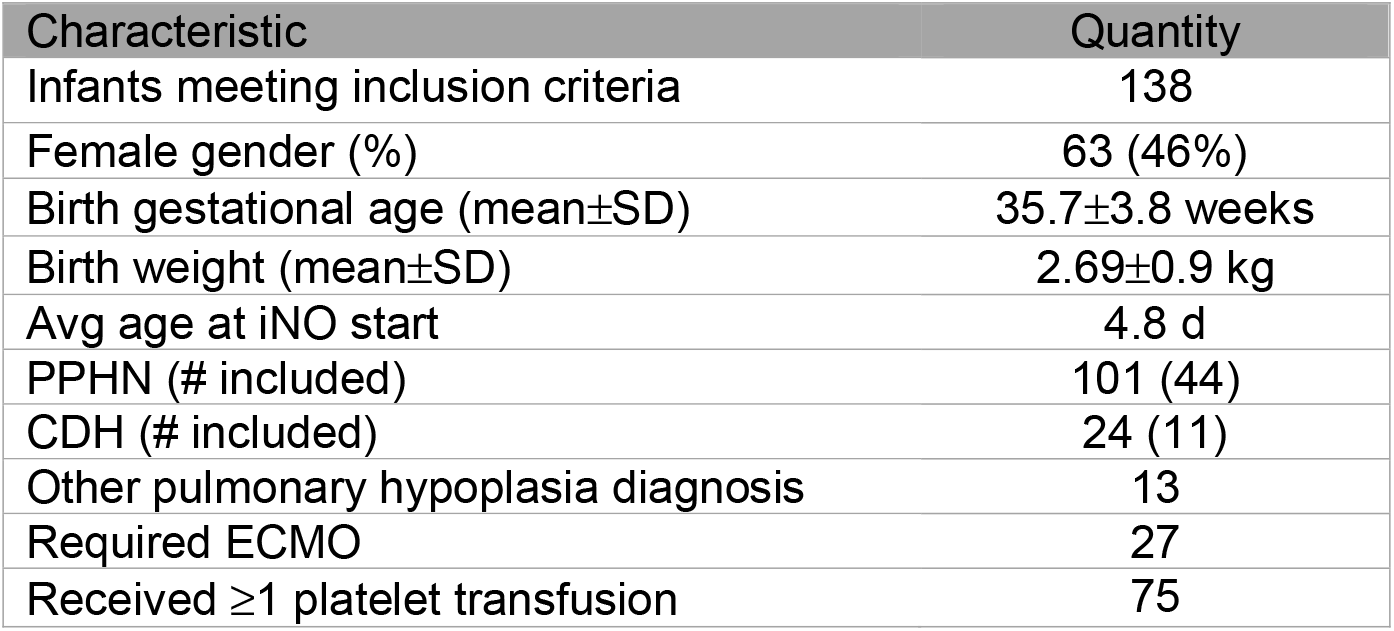
Clinical characteristics of patients that met inclusion criteria for this study. Thrombocytopenia is defined as platelet count <150 ×10^9^ per L. Some patients met multiple exclusion criteria, including need for ECMO or platelet transfusion. Those with ‘other pulmonary hypoplasia’ include diagnosis of omphalocele or idiopathic pulmonary hypoplasia.

We began by assessing platelet count surrounding iNO initiation. Platelet count did not change in the setting of iNO initiation (Fig. 1A, ‘Pre’ vs ‘Start’), despite a significant decrease in respiratory severity scores reflecting improved pulmonary vascular dynamics upon iNO initiation (Fig. 1B). Platelet counts remained stable yet mildly depressed for the duration of iNO therapy.

**Figure 1.**
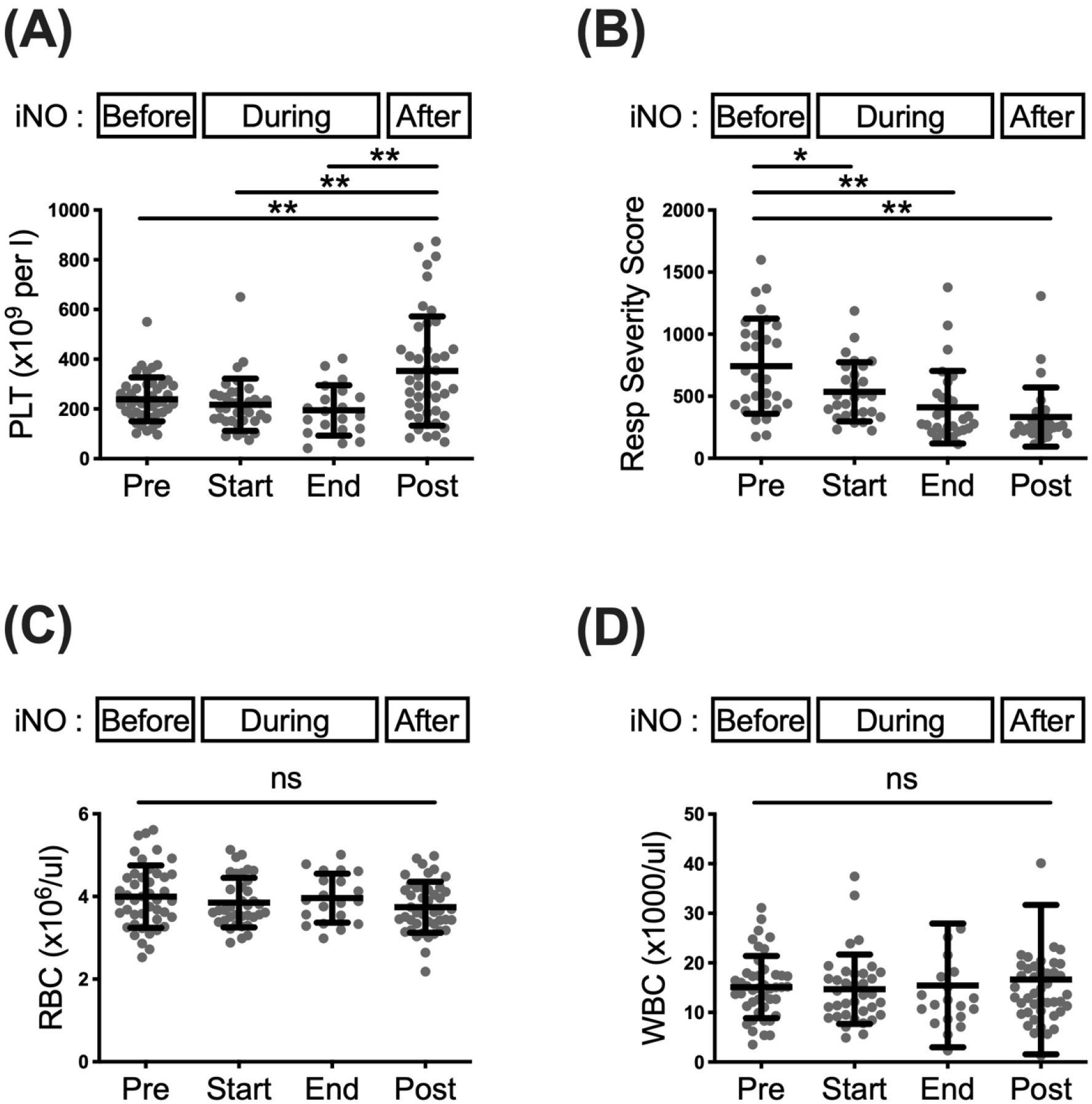
Platelet count in infants with PPHN (excluding CDH or pulmonary hypoplasia) is consistent before and during iNO therapy, but increase after stopping iNO. (A) Platelet counts are relatively consistent before (Pre) and during iNO therapy (Start and End), but acutely increase in the 48 h after stopping iNO (Post). (B) Respiratory severity score significantly decreases following iNO initiation. (C) Red blood cell (RBC) and (D) white blood cell (WBC) counts are consistent in the time periods before, during and after iNO therapy. *p<0.05, **p<0.01. ns, not statistically significant.

Unexpectedly, upon iNO discontinuation there was an acute increase in platelet count in this cohort (Fig. 1A, ‘Post’). We were surprised by the magnitude of change, with platelet count increasing by 50% or an average of 114 ×10^9^ per L compared to pre-iNO treatment levels (239±89 pre-iNO vs 353±220 ×10^9^ per L post-iNO initiation, mean±SD). As comparators, white blood cell (WBC) and red blood cell (RBC) counts remained stable during all analyzed time periods (Fig. 1C-D).

The observed trends in platelet counts, as well as respiratory severity scores and other blood cell counts, were consistent in those with CDH (Supplementary Fig. 1). Of note, there was a trend toward increased platelet count toward the end of iNO therapy that did not reach statistical significance (Supplementary Fig. 1A).

Temporal analyses confirmed that there were no clinically meaningful changes in platelet count in the first 7 d of iNO treatment for these patient cohorts (Supplementary Fig. 2). At minimum, these analyses confirm that platelet count does not acutely increase at the onset of pulmonary vascular dilation in the setting of iNO initiation.

## Discussion

Thrombocytopenia (platelet count <150 ×10^9^ per L) is frequently encountered in the NICU (Gunnink *et al*, 2014) and has been associated with decreased pulmonary blood flow (Mojadidi *et al*, 2014; Le *et al*, 2019). Allogeneic platelet transfusions are standard therapy to increase platelet count, but increase morbidity and mortality in NICU patients (Curley *et al*, 2018).Pulmonary hypertension of is also frequently encountered in the NICU, and large randomized controlled trials have demonstrated the efficacy of iNO in facilitating pulmonary vasodilation (Barrington *et al*, 2017). These studies have not focused on platelet count trends in the setting of PH or iNO therapy.

Of patients meeting inclusion criteria for our study, 37% (31/84) had thrombocytopenia prior to iNO treatment, and almost half (46%) received a platelet transfusion. Infants who received transfusions had significantly lower platelet counts prior to iNO initiation (152±94 ×10^9^ per L, mean±SD) compared with those who did not receive transfusions (239±89 ×10^9^ per L). Hence, neonates with pulmonary hypertension are at significant potential risk for complications associated with thrombocytopenia and its treatment.

Bone marrow-derived platelets take about 7 days to form from stem cells, whereas pre-formed megakaryocytes in the lung might be expected to yield platelets substantially faster. In our cohort, platelet counts did not significantly change following iNO initiation. It is possible that these findings were confounded by dilutional changes related to fluid resuscitation in these critically ill patients, but we would have expected RBCs and WBCs to also fall if this were the case. Alternatively, even minimal pulmonary blood flow in infants with PH might be sufficient to promote platelet elaboration from lung megakaryocytes. Overall, however, bone marrow megakaryopoiesis and thrombopoiesis seem primarily responsible for increased platelet counts seen over time for infants with normal or abnormal lung parenchyma.

We were surprised to see a marked increase in platelet counts following iNO termination. The observed platelet count increase was clinically significant, in similar magnitude to increases seen following 10-15 ml/kg platelet transfusions (Kline *et al*, 2008). Despite historical concern surrounding reduced platelet function in the context of iNO, there has been no correlation between iNO therapy and bleeding (Barrington *et al*, 2017). Biochemical studies suggest that nitric oxide may acutely increase platelet production by promoting megakaryocyte apoptotic mechanisms (Battinelli *et al*, 2002), but over time this might be expected to cause an overall decrease in megakaryopoiesis and/or thrombopoiesis. It is also possible that this change reflects a reactive thrombocytosis following resolution of lung injury, as seen in patients with sickle cell disease (Villagra *et al*, 2007). Further studies are needed to address specific iNO effects that could alter thrombopoiesis in our patient cohort.

In summary, PPHN predisposed our cohort to relative thrombocytopenia that improved once iNO therapy was discontinued. The lack of correlation between pulmonary blood flow and platelet counts suggests that lung-resident megakaryocytes are not a significant untapped reservoir for platelets for this population, at least in the context of PPHN and/or critical illness. However, acute platelet count increases might be expected following iNO discontinuation for some infants. These findings could ultimately help inform empirical platelet transfusion decisions, but it will be important to investigate and confirm these trends in larger cohorts.

## Data Availability

All data will be made available by the authors upon request.

## Acknowledgement

The authors are grateful for thoughtful suggestions from Amy Padula, PhD, MSc.

## Statement of Ethics

This study was deemed exempt from review by the Children’s Hospital of Philadelphia Institutional Review Board (IRB).

## Authorship Contributions

CST, MPL and MAP designed the study. CST, MD, SK, EAJ, MPL, and MAP collected, analyzed and interpreted data. CST wrote the paper. All authors revised and approved the submitted version of the manuscript.

## Supplementary Figures

**Supplementary Figure 1.**
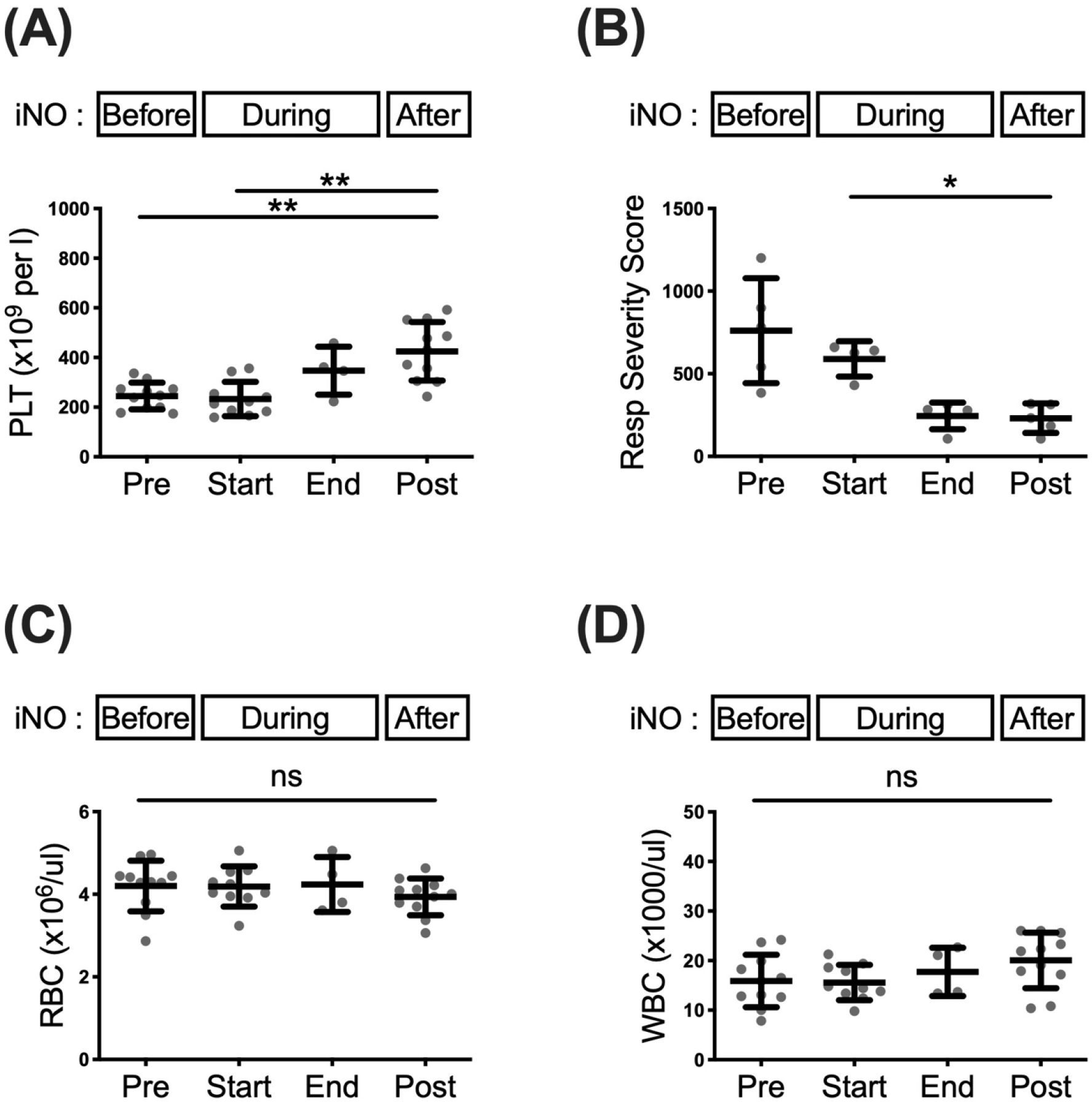
Platelet count in infants with congenital diaphragmatic hernia do not change upon iNO initiation, but increase after stopping iNO. (A) Platelet counts post-iNO cessation significantly increase compared to platelet counts before starting (Pre) or just after starting (Start) iNO. (B) Respiratory severity score significantly decreases after completing iNO therapy. (C) Red blood cell (RBC) and (d) white blood cell (WBC) counts are consistent in the time periods before, during and after iNO therapy. *p<0.05, **p<0.01. ns, not statistically significant.

**Supplementary Figure 2.**
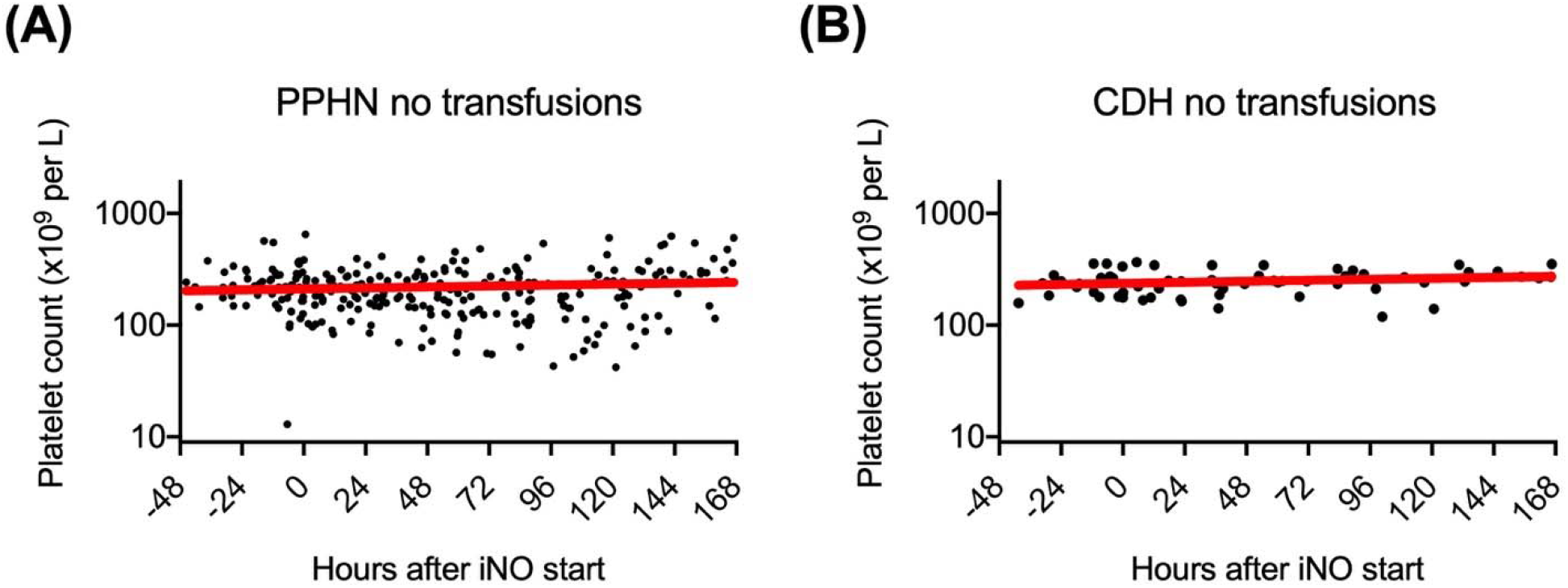
Temporal platelet count trends in infant platelet counts following iNO initiation. Black dots represent platelet counts at the indicated time points. Solid red linear regression lines depict platelet count trends from 48 h before initiating iNO through 168 h (1 week) after initiating iNO for (A) infants with PPHN who did not receive platelet transfusions (slope=0.18, p=0.18, no significant deviation from 0) or (B) infants with CDH who did not receive platelet transfusions (slope=0.22, p=0.11, no significant deviation from 0).

